# Comparing Wastewater-Based and Case-Based *R*_*t*_ Estimates of SARS-CoV-2 Transmission in Georgia, USA Using Generalised Linear Mixed Models

**DOI:** 10.1101/2025.10.24.25338734

**Authors:** Seth Edmunds, Doug Landsittel, Marco Ajelli, Maria Litvinova

## Abstract

The COVID-19 pandemic has highlighted limitations in case-based surveillance due to inconsistent testing and reporting. Wastewater-based epidemiology (WBE) has emerged as a complementary surveillance approach for tracking SARS-CoV-2 transmission, capturing both symptomatic and asymptomatic infections. The aim of this study is to evaluate the effectiveness of WBE in estimating the effective reproduction number (*R*_*t*_) of SARS-CoV-2 in Georgia, USA. We used a Generalised Linear Mixed Model to analyse viral concentration data from multiple wastewater treatment plants (WWTPs) collected between June 1, 2022 and December 15, 2022. After controlling for flow rates and site-level heterogeneity, model residuals were transformed into a non-negative incidence-like series used to estimate wastewater-based *R*_*t*_. Wastewater-based *R*_*t*_ was compared with case-based *R*_*t*_ estimates using Spearman correlation. The two *R*_*t*_ estimates showed concordant temporal patterns across most sites, with stronger correlations in areas with higher case counts (Spearman correlations ranging from 0.39 to 0.84, *p* < 0.001). Wastewater-based *R*_*t*_ tracked increases and decreases in transmission over similar time scales as case-based estimates, while exhibiting reduced sensitivity to short-term changes in clinical testing and reporting behaviour. These findings suggest that WBE can support estimation of transmission trends and complement traditional case-based surveillance for public health monitoring.

## Introduction

The COVID-19 pandemic has underscored the need for effective surveillance systems to monitor disease transmission at the population level. Initially, case-based surveillance was critical for tracking the spread of COVID-19. However, fluctuations in testing rates and the changes in reporting practices gradually undermined its reliability. By late 2022, the shift by the Centers for Disease Control and Prevention (CDC) from daily to weekly aggregated case and death reporting reduced the temporal resolution of data available for public health monitoring. This change was intended to reduce the reporting burden and improve operational efficiency, but it limited the ability to assess localised and short-term transmission dynamics. After the expiration of the public health emergency on May 11, 2023, the U.S. Department of Health and Human Services (HHS) could no longer require laboratories to submit COVID-19 testing results; subsequently, hospital COVID-19 reporting requirements were discontinued effective May 1, 2024 (1). Together, these changes highlight the need for complementary surveillance approaches capable of monitoring population-level transmission when clinical reporting is incomplete or delayed.

In response to these challenges, wastewater-based epidemiology (WBE) has re-emerged as a promising complementary approach, providing a non-invasive, scalable method to track viral RNA in community wastewater, capturing both symptomatic and asymptomatic infections. WBE offers a unique opportunity to supplement traditional case-based surveillance, particularly by providing a population-level signal that is less dependent on individual testing behaviour and can reflect changes in community transmission. Because wastewater surveillance occurs at the community level, it is less sensitive to biases arising from uneven access to testing and underreporting, making WBE especially valuable in settings with limited healthcare access or inconsistent clinical testing (2, 3).

With the rise of at-home COVID-19 testing, which is largely not reported, comparing wastewater-based and clinical estimates of the effective reproduction number (*R*_*t*_) has become increasingly important for understanding transmission dynamics while recognizing that each data stream reflects different processes and sources of spatio-temporal heterogeneity. Recent studies have begun to leverage wastewater data alongside clinical surveillance to infer transmission dynamics and develop early-warning systems under varying assumptions about shedding, reporting, and observation processes (4). By capturing SARS-CoV-2 RNA at the community level, wastewater-based epidemiology (WBE) provides a population-scale signal that is less influenced by biases in clinical data, such as uneven testing access, changing individual behaviour, or reporting delays, though it is subject to its own sources of variability (5

Several studies have estimated wastewater-based SARS-CoV-2 effective reproduction numbers using various assumptions, preprocessing, and model complexities (2, 6) While county-level estimations have shown high concordance with traditional case-based estimates (7), there is a need to validate these methods at finer scales. Using a unique dataset that includes both reported cases and wastewater surveillance at the level of individual sewersheds, we demonstrate the concordance of *R*_*t*_ estimations on a disaggregated level. This study provides evidence for the operational utility of wastewater-based *R*_*t*_ estimations and offers a practical pipeline accounting for sewershed-related heterogeneity.

The aim of this study is to evaluate the effectiveness of WBE in estimating *R*_*t*_ of SARS-CoV-2 at a sewershed level. We focus on data collected between June 1 and December 15, 2022 - a period during which routine case reporting via electronic laboratory and provider reports remained available, enabling a contemporaneous comparison of wastewater-based and case-based *R*_*t*_ estimates. To compare these signals, we apply a Generalised Linear Mixed Model (GLMM) to wastewater concentrations, adjusting for temporal trends, flow-related dilution, and persistent sewershed differences. This approach allows us to assess the degree to which wastewater-derived transmission dynamics align with contemporaneous case-based *R*_*t*_ estimates at the sewershed level, and to evaluate the role of WBE as a complementary surveillance signal for clinical data at finer scales. (1, 8).

## Methods

### Data Collection

#### Wastewater Data Sources

WWTP employees collected twice-weekly wastewater samples, which were shipped to two university laboratories for analysis. Sampling frequency was typically twice weekly across sites, though exact schedules varied by facility and operational constraints (summarized in Appendix Table A1). Viral RNA concentrations were quantified using PCR, specifically targeting the N2 gene for its widespread use and comparable performance to the N1 target (Huisman et al., 2022). RT-qPCR was used at WWTP 9 following the method detailed by Lott et al. (2023) due to logistical constraints, while dPCR, preferred for its higher sensitivity to low viral concentrations, was used at all other sites following the protocol described by Sablon et al. (2022) (Sablon et al. 2022). Differences in analytical platform across sites were treated as time-invariant site characteristics and accounted for through site-level random effects in downstream modelling. Flow rate data were obtained from routine facility records and aligned with wastewater sampling dates; depending on facility practice, measurements corresponded to composite (time- or flow-weighted) or grab sampling approaches, consistent with standard wastewater surveillance reporting practices (11). Concurrent flow rates were incorporated to account for dilution-related variability in wastewater volume.

#### Case Data Sources

Case data were obtained from routine provider and electronic laboratory reports submitted to the Department of Public Health (DPH) and stored within an electronic disease reporting system. Residential addresses were geocoded to obtain XY coordinates. Sewershed shapefiles were provided by the facilities. Case-to-sewershed assignment assumes accurate residential geocoding and alignment with sewershed boundaries, consistent with standard public health surveillance practice. Case data were aggregated daily by sewershed geography using symptom onset date when available; specimen collection date was used otherwise.

### Data Processing

#### Case Data Processing

Cases were assigned to sewersheds based on geocoded residential addresses and the spatial overlap with sewershed boundaries. Epidemic curves were generated using either the date of symptom onset or, if unavailable, the date of the first positive specimen (including asymptomatic cases). Missing daily case counts were imputed using Kalman filtering to provide smoothed estimates, ensuring the preservation of temporal sequences while handling reporting fluctuations. Imputation primarily reflects completion of a daily time series from routine non-daily reporting rather than filling extended gaps in surveillance and the Kalman approach was selected based on comparative performance in preserving temporal structure, as summarized in the Appendix. Data were restricted to the study period from June 1, 2022, to December 15, 2022, and the processed case data were used in conjunction with wastewater viral concentrations for analysis.

#### Wastewater Data Processing

All data are from 8 sewersheds in Georgia, USA. Outliers in wastewater viral concentrations and flow rates were detected using the Interquartile Range (IQR) method, with outliers replaced by the median to reduce their influence on downstream analysis. Flagged values were treated as transient spikes for robustness rather than assumed measurement errors, and the number of flagged observations by site is reported in Appendix Table A2. While Appendix Table A1 summarizes the characteristics of all 10 facilities participating in the broader state surveillance program, two facilities, WWTP 4 and WWTP 7, lacked sufficient data during the study period window (June 1 – December 15, 2022). Because only observations exceeding the IQR threshold were modified, minimum and maximum values could remain unchanged if those extremes did not meet the outlier criterion. This method was selected based on a sensitivity analysis comparing multiple outlier detection techniques, including Median Absolute Deviation (MAD), using agreement with case-based *R*_*t*_ (Spearman correlation and RMSE) as evaluation criteria. Samples were predominantly 24-hour composite, collected twice weekly. Population served by individual sewersheds varied from 10,000 (WWTP 10) to 650,000 (WWTP 1) Sewershed locations spanned urban, suburban, and rural areas across Georgia; specific geographic coordinates are not disclosed to protect facility identities, but site-level population, flow, and sample characteristics are summarized in Appendix Table A1. Concentrations were not pre-normalised. Instead, by including flow as a fixed effect in the GLMM, the model inherently adjusts for flow-related dilution, with the resulting residuals representing viral signal fluctuations independent of flow variation.

Preprocessing method selection was guided by comparative evaluation across multiple approaches. Outlier detection using the IQR method flagged 27 concentration values and 57 flow values across sites, compared with 27, and 123 respectively for the MAD approach. Missing data imputation using Kalman filtering (71.8% of the daily time series) reflected completion to daily resolution from approximately twice-weekly sampling; inter-sample gaps had a median of 2 days. Sensitivity analyses across rolling window lengths showed that 14-day windows achieved the highest concordance with case-based *R*_*t*_ estimates (median Spearman ρ = 0.608, IQR = 0.121), comparable to 10-day windows (ρ = 0.612, IQR = 0.184) and superior to 7-day windows (ρ = 0.593, IQR = 0.254). The selected preprocessing combination (IQR outlier detection, Kalman imputation, 14-day rolling average) balanced noise reduction with preservation of transmission trends and minimized data loss (flagging fewer than half as many flow outliers as the MAD approach), as assessed by correlation with case-based estimates (median RMSE = 0.098). Complete site-level characteristics, outlier counts, missingness patterns, and sensitivity results are provided in Appendix Tables A1-A4.

Missing data were interpolated using Kalman filtering, chosen for its performance relative to linear and spline interpolation methods in preserving temporal structure during routine non-daily sampling. (Miyazawa et al. 2024; Narci et al. 2021) Both viral concentrations and flow rates were log-transformed to stabilise variance and normalise the data distribution. Comparisons of raw and processed data are presented in Appendix Table A5. These preprocessing steps were implemented to stabilise the time series for downstream transmission trend estimation rather than to reconstruct true incidence or shedding processes.

### Statistical Modelling

#### Model Specification

A Generalised Linear Mixed Model (GLMM) was employed (significance level set to α=0.05) to analyse viral concentrations in wastewater, with both fixed and random effects to capture the hierarchical and temporal structure of the data. The fixed effects included time and flow rate, while the random effects accounted for site-level variability across WWTPs. Model diagnostics were used to assess statistical fit and stability of the mixed-effects model, including residual behaviour and random-effect variance. While these diagnostics ensure the statistical robustness of the framework, they do not, by themselves, imply the biological validity of the resulting residual time series as a direct and comprehensive measure of the number of infections. By accounting for both site-specific and time-based variability, the model provided a flexible framework for analyzing transmission dynamics signal. (14, 15)

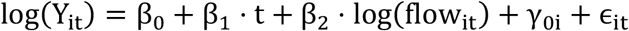

Where:

- *Y*_*it*_ is the viral concentration in wastewater at time *t*for WWTP *i*.
- β_0_ is the fixed intercept representing the global baseline concentration.
- β_1_ is the coefficient for the linear temporal trend.
- β_2_ is the coefficient adjusting for the log-transformed wastewater flow rate.
- *t* is a daily time variable to account for temporal trends.
- flow_it_ is the flow rate of wastewater at time *t* for WWTP *i*.
- γ_0i_ is the random intercept for WWTP *i*, capturing time-invariant site-specific differences where 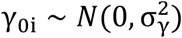.
- *ϵ*_*it*_ is the residual error term.

The primary specification includes a fixed effect for temporal trend and a site-specific random intercept, implying a common temporal trend with persistent site-specific differences. By utilizing a GLMM instead of manual flow-normalization, we provide a unified framework to adjust simultaneously for flow-related dilution, shared temporal trends, and site-level heterogeneity (e.g., variations in measurement and sampling approaches) that are captured by the random intercept (γ_0i_). By intentionally omitting site-specific random slopes for time, we ensure that localised deviations from the regional trend are preserved within the residuals (ϵ_it_) rather than being absorbed into the model parameters. This approach allows the residuals to more accurately reflect the site-level dynamics necessary for subsequent R_t_ estimations while accommodating the nested data structure.

It is important to note that the linear time covariate is specified as a global fixed effect across a geographically diverse set of sewersheds. It is intended to control for shared baseline trend without absorbing localised epidemic waves. However, researchers applying this framework should note that if applied to highly synchronized, homogeneous sites over a short period, the global time effect may absorb the epidemic signal, especially during the period of consistent epidemic growth or decline. In such cases, the approach should be implemented without the time trend fixed effects. Empirical demonstrations of the model’s robustness across varying observation windows and distinct transmission phases are provided in Appendix Figures A1 and A2, respectively.

#### Residual Extraction and Smoothing

Residuals ϵ_it_ from the GLMM were extracted to capture variability in viral concentrations not explained by the fixed effects of time, flow rate, and site-specific differences in wastewater concentrations, including measurement differences. The inclusion of site-specific random intercepts accounts for persistent between-site differences in baseline viral concentration, allowing within-site temporal deviations to be examined on a comparable scale. To reduce high-frequency noise, a 14-day rolling average was applied to the residual series. While rolling averages introduce additional temporal smoothing and may attenuate short-term changes, this window length reflects a pragmatic balance between noise reduction and trend interpretability and was evaluated through sensitivity analyses reported in Appendix Table A4. This approach ensures that critical transmission patterns are preserved while minimizing the impact of daily fluctuations. Accordingly, residual-based analyses are intended to support inference on transmission trends rather than to reconstruct individual-level infection, shedding processes, case or hospitalization incidence.

### Estimation of the Effective Reproduction Number (Rt)

#### Rt Calculation

The effective reproduction number (*R*_*t*_) was estimated using the EpiEstim package in R. (16, 17) A parametric serial interval distribution was specified with a mean of 2.75 days and a standard deviation of 2.53 days, utilizing updated parameters specific to the SARS-CoV-2 Omicron variant. (Kremer et al., 2022) To align with the requirements for *R*_*t*_ estimation, the GLMM residuals were exponentiated to back-transform the values from the log-link model and ensure the non-negativity required for an incidence proxy. Two estimates of *R*_*t*_ were calculated: one based on case-derived incidence, and one based on a transformed wastewater-derived residual series. Both case-based and wastewater-based series were smoothed using a 14-day rolling average prior to *R*_*t*_ estimation to reduce short-term noise, with the understanding that this additional smoothing further convolves the underlying signal; the impact of window length on *R*_*t*_ concordance was evaluated through sensitivity analyses reported in the Appendix Table A4. (2, 3) As a result, wastewater-derived *R*_*t*_ estimates are interpreted as a signal of relative transmission dynamics rather than as direct estimates of intrinsic incidence-based reproduction parameters.

#### Sensitivity Analysis: Rolling Average Window

A sensitivity analysis was performed to assess the impact of rolling average window length on both case-based and wastewater-based *R*_*t*_ estimates. Rolling windows ranging from 3 to 30 days were evaluated, with performance assessed based on agreement with case-based *R*_*t*_ (Spearman correlation and RMSE) and qualitative stability of inferred trends. The 14-day window was selected for primary analysis because it provided a pragmatic balance between noise reduction and temporal responsiveness across sites, as summarized in Appendix Table A4. Longer windows further reduced noise but increasingly attenuated short-term changes during periods of rapid transmission change. (19)

#### Estimate Alignment

To quantify concordance between wastewater-based and case-based *R*_*t*_ estimates, we computed Spearman rank correlation coefficients across sites. Analyses were restricted to the period prior to December 2022, after which changes in case reporting requirements substantially altered the reliability and completeness of case-based surveillance. Alignment was further assessed by comparing the temporal co-movement of wastewater-based *R*_*t*_, case-based *R*_*t*_, and reported case counts, focusing on direction and timing of major transmission changes rather than pointwise equality. This approach emphasizes concordance in transmission trends under differing observation processes rather than validation against a presumed ground truth. (20)

## Results

The initial dataset comprised 294 observations for both PCR target average concentration and flow rate measurements across eight WWTPs. After data preprocessing, which included outlier handling and missing data imputation, the number of observations increased to 1,019. Detailed statistics on preprocessing outcomes are presented in the Appendix. A total of 749 data points (72% of the dataset) were imputed using Kalman smoothing reflecting routine completion of a daily time series from non-daily (typically twice-weekly) sampling rather than extended gaps in surveillance. Kalman smoothing was employed because it effectively estimates missing values in time series data by modelling the underlying stochastic processes. This method accounts for variability by incorporating both the observed data and the process noise, allowing for smoothed estimates of the underlying temporal signal amidst measurement uncertainty and routine sampling variability. (Narci et al. 2021)

An overview of the *R*_*t*_ estimates derived from both case-based and wastewater-based surveillance is provided in Table 1. This table includes the duration of data collection, total case counts, median *R*_*t*_ values, and Spearman correlation coefficients across the eight wastewater treatment plants (WWTPs). All statistically significant results illustrate notable positive correlation between the two *R*_*t*_ estimations, with the highest correlation being observed for WWTP 5 (0.84, *p* < 0.001) and the lowest for WWTP 3 (0.39, *p* < 0.001). The results (Table 1) indicate that average case burden influenced both correlation strength and statistical significance (Figure 1). For example, no statistically significant correlation was observed for WWTP 9; while its data collection duration was comparable to WWTP 1, its average case burden was substantially lower. Generally, correlation strength and statistical significance aligned with overall case burden. Sewersheds with very low daily case counts frequently lacked the temporal variability required to power rank-based correlation (e.g., WWTP 9 and WWTP 6). However, statistically significant correlations were occasionally observed even in low-incidence settings (e.g., WWTP 10, averaging approximately 2 cases per day). The overall pattern is illustrated in Figure 1, where *R*_*t*_ concordance generally increases as a function of the average number of cases.

**Table 1.** Core Statistics of *R*_*t*_ Estimates, Correlations and Sewershed Characteristics (ordered by descending case burden)

| Sewershed | Duration (days) | Total Cases | Median $R_t$ (Case) | Median $R_t$ (WW) | Spearman Correlation | P-Value |
| --- | --- | --- | --- | --- | --- | --- |
| WWTP 1 | 191 | 15288 | 1 | 1 | 0.73 | <0.001 |
| WWTP 3 | 191 | 2564 | 1 | 1 | 0.39 | <0.001 |
| WWTP 5 | 177 | 1560 | 1.02 | 1.03 | 0.84 | <0.001 |
| WWTP 8 | 191 | 1100 | 1.02 | 1.03 | 0.58 | <0.001 |
| WWTP 2 | 26 | 952 | 1.11 | 1.17 | 0.95 | <0.001 |
| WWTP 9 | 191 | 503 | 1.06 | 1.02 | -0.01 | 0.841 |
| WWTP 6 | 26 | 126 | 1.06 | 1.23 | 0.38 | 0.052 |
| WWTP 10 | 26 | 52 | 1.06 | 1.2 | 0.41 | 0.035 |

**Figure 1.**
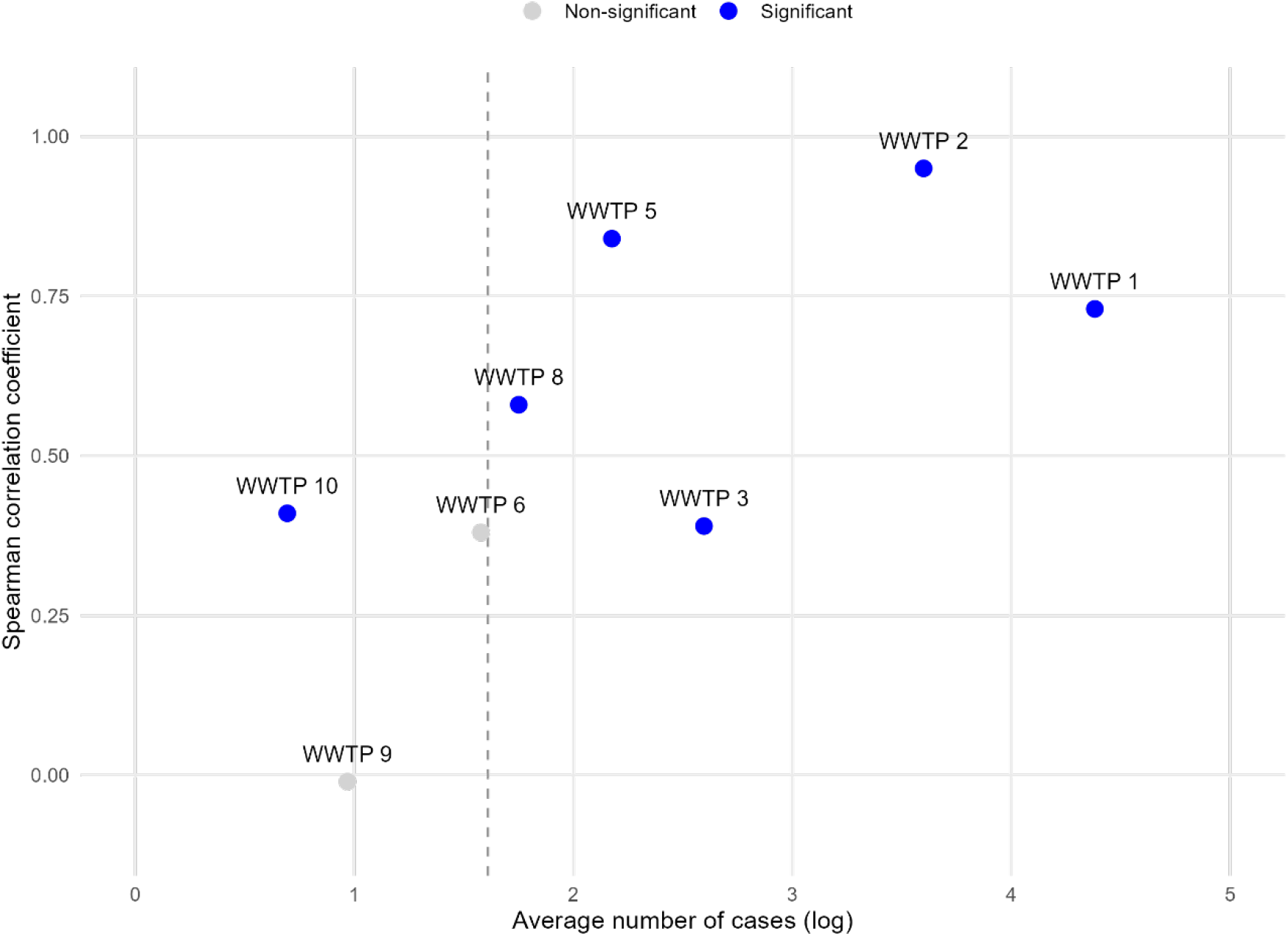
Average Number of Cases and Spearman Correlation Coefficient Between Case-Based and Wastewater-Based *R*_*t*_ Estimates by Wastewater Treatment Plant Sewershed The vertical dashed line indicates approximately five average daily cases (log scale), below which rank-based correlation was generally underpowered in our data, though exceptions were observed (e.g., WWTP 10).

**Figure 2.**
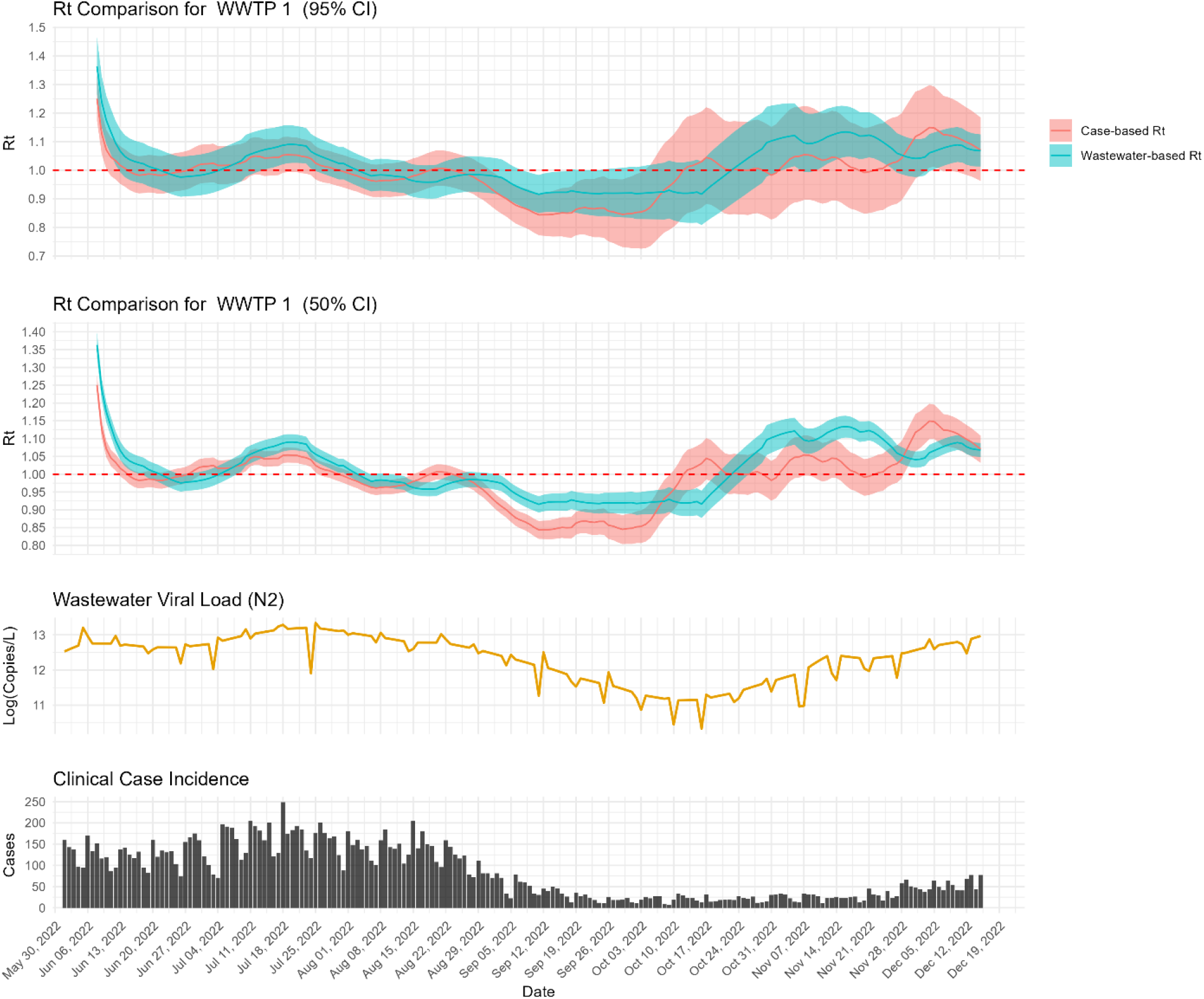
Comparison of Wastewater-Based and Case-Based *R*_*t*_ Estimates for WWTP 1 At WWTP 1, the wastewater-based and case-based *R*_*t*_ estimates closely tracked from June through August, both peaking in mid-July, which aligns with the peaks in the interpolated wastewater signal and clinical case incidence (Figure 2). By late August, both estimates fell below the epidemic threshold (Rt=1). From mid-October onward, the wastewater-based climbed above epidemic threshold earlier than the case-based estimate in this sewershed.

**Figure 3.**
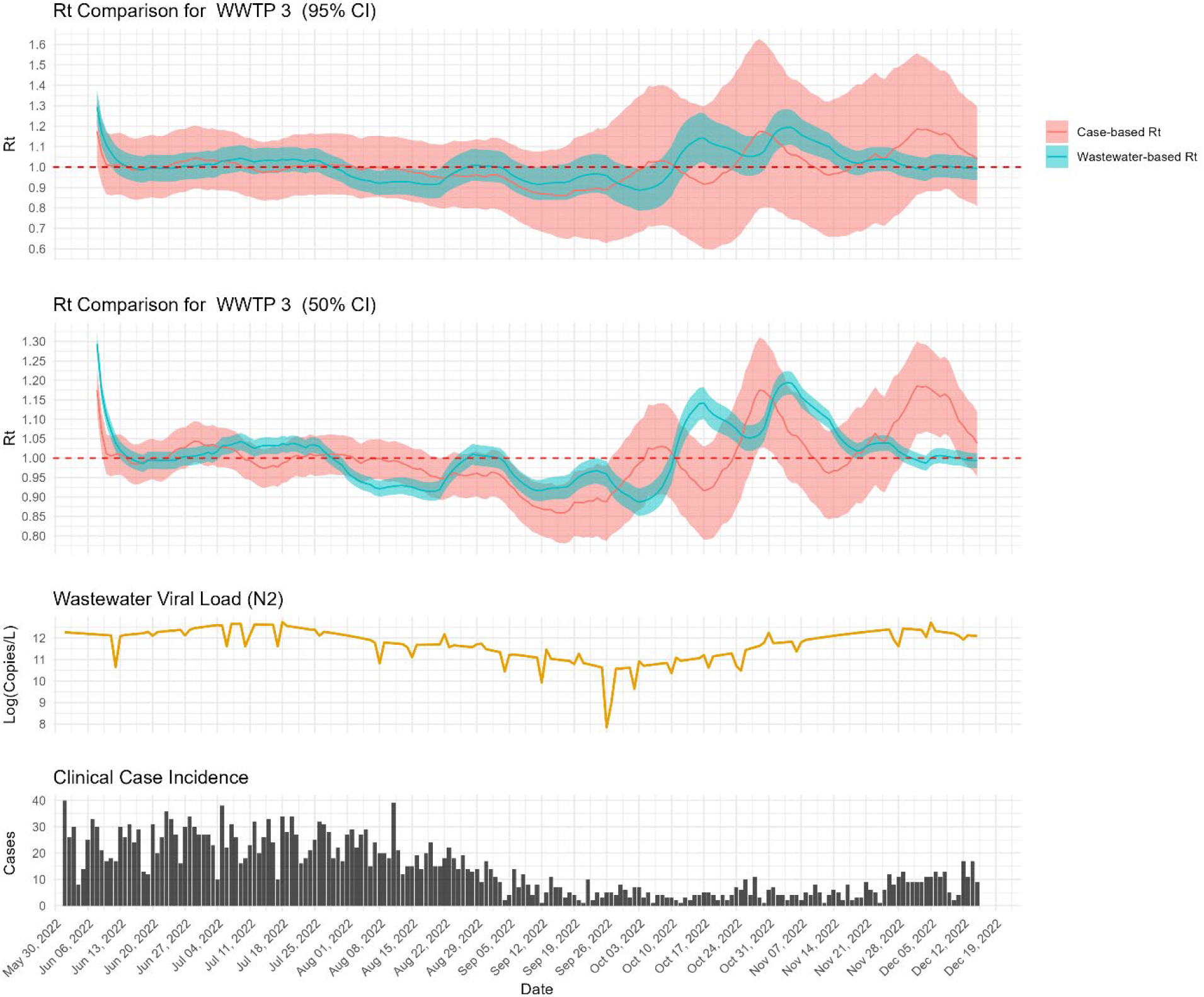
Comparison of Wastewater-Based and Case-Based *R*_*t*_ Estimates for WWTP 3 In WWTP 3, the wastewater-based estimates sometimes lagged peaks in the case-based *R*_*t*_ (Figure 3). In mid-October, the wastewater-based *R*_*t*_ increased to approximately 1.1−1.3, with comparatively narrower confidence intervals, while the case-based estimates exhibited wider uncertainty that encompassed the threshold of 1.0.

**Figure 4.**
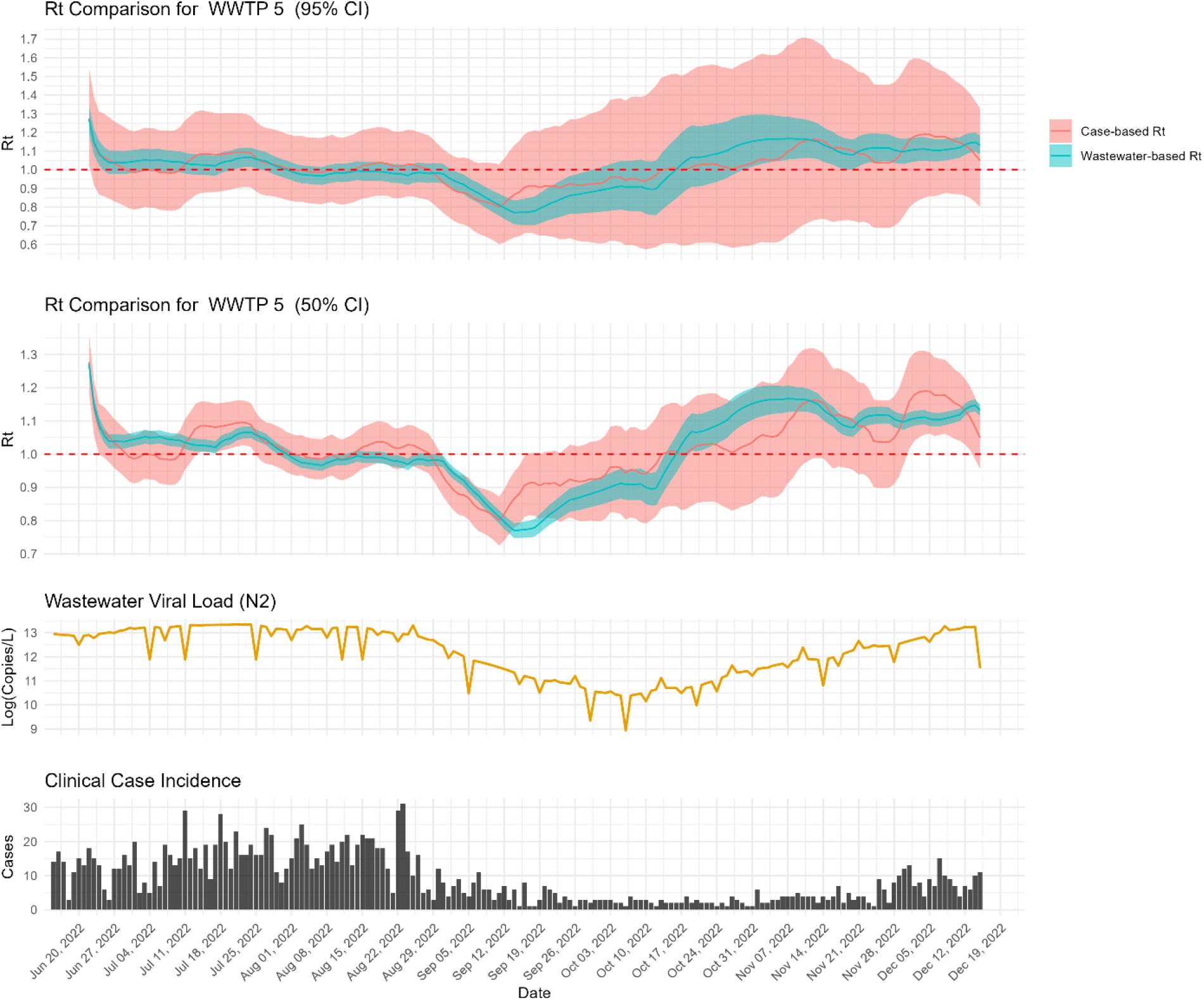
Comparison of Wastewater-Based and Case-Based *R*_*t*_ Estimates for WWTP 5 For WWTP 5, the wastewater-based *R*_*t*_ initially aligned with case-based trends during early summer, with values above 1.0 (Figure 4). In mid-September, the wastewater-based *R*_*t*_ declined below 0.8 prior to a similar decline in the case-based estimate. A subsequent increase in the wastewater-based *R*_*t*_ in mid-October preceded a rise in cases.

**Figure 5.**
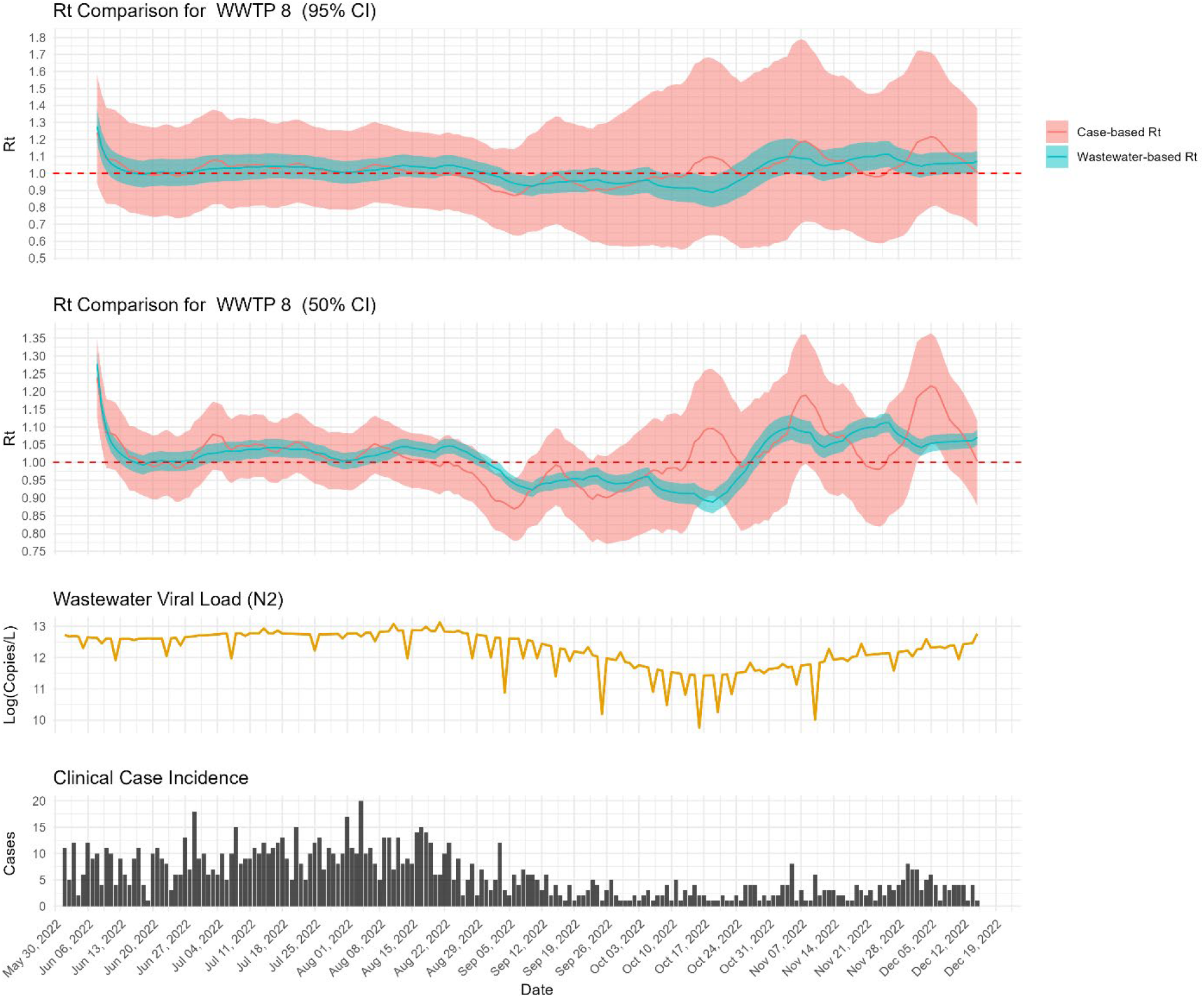
Comparison of Wastewater-Based and Case-Based *R*_*t*_ Estimates for WWTP 8 At WWTP 8, the wastewater-based *R*_*t*_ remained above 1.0 from early July, with brief dips (Figure 5). Case-based *R*_*t*_ exhibited broadly similar temporal patterns but with shifts in timing consistent with differences in smoothing and observation processes.

**Figure 6.**
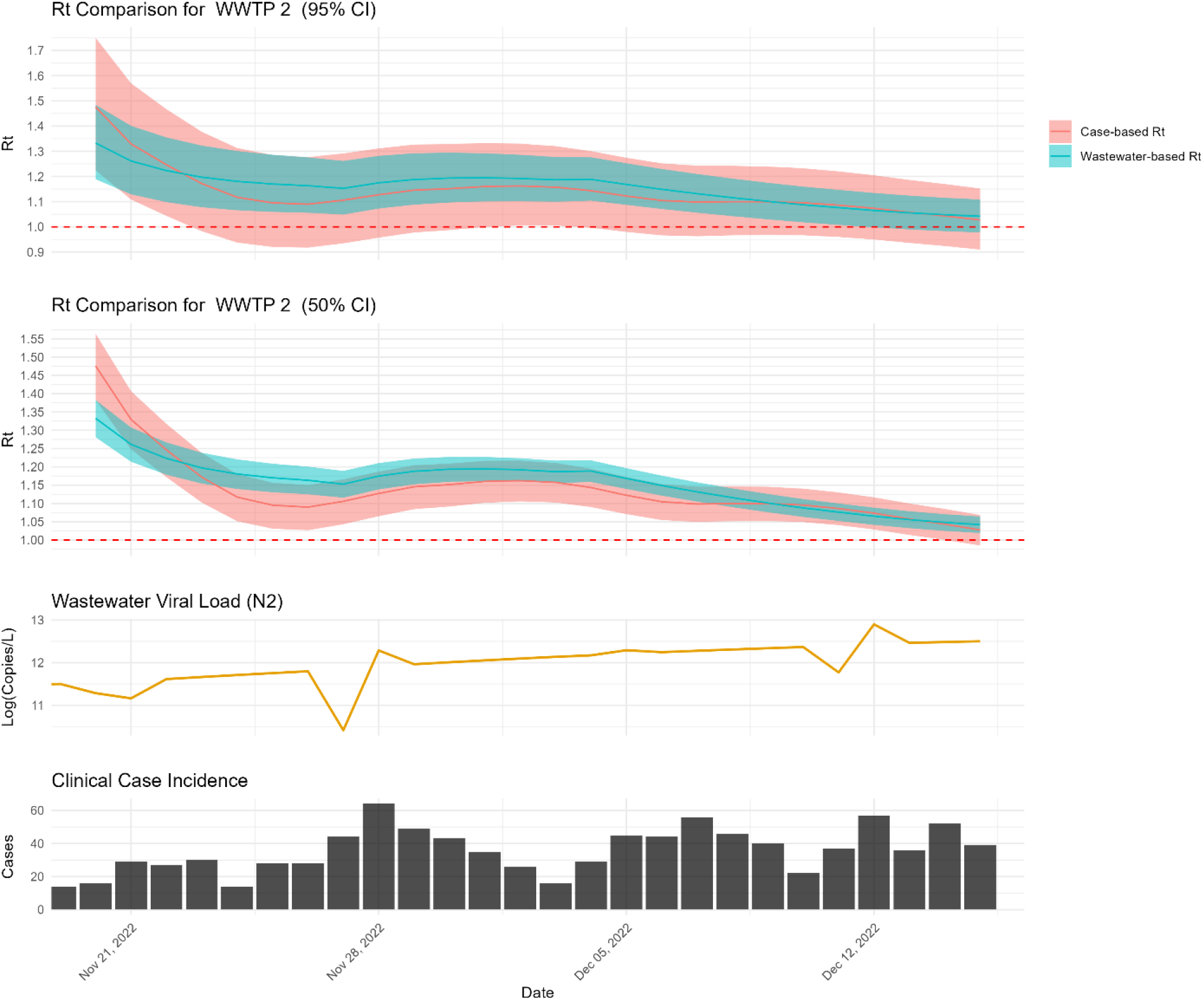
Comparison of Wastewater-Based and Case-Based *R*_*t*_ Estimates for WWTP 2 At WWTP 2, over the shorter study period, the wastewater-based *R*_*t*_ remained consistently above 1.0, while the case-based *R*_*t*_ rose above 1.0 in early December (Figure 6). An initial elevation in the wastewater-based estimate was interpreted as a transient anomaly consistent with preprocessing and smoothing effects rather than sustained transmission change.

### Overview of Rt Estimates Across All Sites

Table 2. Percent of Days with *R*_*t*_ Confidence Intervals Above or Below 1 (ordered by descending case burden)

**Table 2.** illustrates the percentage of days on which lower and upper bounds of the *R*_*t*_ confidence intervals (CIs) were either above or below the epidemic threshold (1.0). This metric provides supplementary information on directional agreement relative to the epidemic threshold (*R*_*t*_ = 1), but was not used as a primary performance criterion because it is sensitive to smoothing choices and uncertainty width.

| Sewershed | Case-Based |  |  |  | Wastewater-Based |  |  |  |
| --- | --- | --- | --- | --- | --- | --- | --- | --- |
|  | 95% CI |  | 50% CI |  | 95% CI |  | 50% CI |  |
|  | > 1 | < 1 | > 1 | < 1 | > 1 | < 1 | > 1 | < 1 |
| WWTP 1 | 3.14 | 17.28 | 20.94 | 28.8 | 25.13 | 7.85 | 43.46 | 33.51 |
| WWTP 3 | 0.52 | 0 | 9.95 | 12.04 | 12.57 | 12.57 | 37.7 | 30.89 |
| WWTP 5 | 0.56 | 0 | 14.69 | 10.17 | 31.64 | 15.82 | 54.24 | 33.9 |
| WWTP 8 | 0 | 0 | 4.71 | 4.19 | 8.9 | 8.38 | 49.74 | 27.75 |
| WWTP 2 | 19.23 | 0 | 96.15 | 0 | 84.62 | 0 | 100 | 0 |
| WWTP 9 | 0 | 0 | 8.38 | 0 | 2.09 | 0 | 40.84 | 8.38 |
| WWTP 6 | 0 | 0 | 7.69 | 0 | 88.46 | 0 | 96.15 | 0 |
| WWTP 10 | 0 | 0 | 7.69 | 0 | 65.38 | 15.38 | 73.08 | 19.23 |

### Detailed Analysis of Representative Sites

The wastewater-based and case-based *R*_*t*_ estimates were analysed across five representative sites with total case incidence greater than 1,000 cases during the study period. Each site exhibits unique patterns in transmission dynamics, as captured by the two surveillance methods. Sites were selected to represent higher-incidence sewersheds where case-based *R*_*t*_ estimates showed sufficient variability to support meaningful temporal comparisons. The sites are presented in the order of decreasing case burden.

Two levels of uncertainty around median estimates (50% and 95% Cis) are used to represent two levels of potential risk-tolerance of public health practitioners in making decisions based on uncertain data.

## Discussion

Our analyses indicate that wastewater-based *R*_*t*_ estimates exhibited broadly similar temporal patterns to case-based estimates across most sites, capturing transmission shifts within approximately two weeks of clinical data. The fundamental purpose of evaluating a clinical comparator is to determine whether pathogen detection in wastewater translates into a tangible public health burden. However, selecting an appropriate clinical comparator requires navigating the inherent limitations of all potential public health signals. Because *R*_*t*_ estimation relies on the ratio of successive incidence values rather than absolute counts, it remains mathematically robust to systematic underreporting provided ascertainment does not shift abruptly. Therefore, the optimal choice must be guided by the stability of the signal during the observation window and the specific practical needs of public health authorities.

For this study, we utilized reported case incidence by symptom onset or collection date. While such data inherently underestimates true infection burden, as demonstrated by infection incidence being 2.2 to 3.7 times higher than reported cases in the United States during the Omicron wave (21), reporting requirements during our study period (June– December 2022) were sufficiently stable to establish a reliable baseline. The subsequent expiration of public health mandates and the wider utilization of non-reportable at-home testing rapidly rendered clinical ascertainment lower and highly volatile (21), necessitating our choice of the study’s end date. Therefore, for this study, reported cases provided the most stable, geospatially appropriate comparator to address the local public health burden at the sewershed level. However, during periods of rapid surveillance infrastructure changes, divergence between wastewater and clinical signals should not be interpreted as a failure of wastewater surveillance, but rather as a complementary reflection of community transmission dynamics essential for comprehensive situational awareness.

Several limitations should be acknowledged. A substantial portion of the time series required completion to daily resolution for *R*_*t*_ estimation, primarily reflecting routine non-daily sampling rather than prolonged data gaps. Kalman smoothing was used to interpolate missing days and stabilise temporal structure, which may attenuate short-term variability but is necessary for renewal-based estimation frameworks. Additionally, wastewater measurements reflect a convolution of infections over heterogeneous shedding durations and sewer transport processes. As a result, wastewater-derived *R*_*t*_ should be interpreted as a smoothed indicator of transmission trends rather than a direct measure of incident infections. This interpretation is subject to uncertainties and heterogeneities in SARS-CoV-2 shedding likelihood, rates, duration, and routes.

Furthermore, while we account for the flow differences, variability introduced by site-specific shedding dynamics, viral variants, and in-sewer environmental factors impacts the precision of these estimates. Nevertheless, these uncertainties do not preclude the detection of major transmission shifts as evidenced by the strong concordance observed with case data in higher-burden areas.

Finally, assignment of COVID-19 cases to sewersheds relied on residential address geocoding, which may misclassify infections if substantial exposure or shedding occurred outside the residential sewershed. Future studies could enhance geolocation accuracy by incorporating additional data sources or methods, such as improved address resolution or alternative approaches to case-to-sewershed assignment. These limitations highlight areas for methodological improvement but do not alter the central finding that wastewater-based *R*_*t*_ estimates captured transmission trends broadly consistent with case-based surveillance. Rather than enabling direct intervention decisions, wastewater-derived *R*_*t*_ provides a complementary indicator of changing transmission dynamics that may support situational awareness when clinical surveillance is incomplete or delayed.

The weaker correlation observed at WWTP 3 (ρ = 0.39) despite higher total case counts compared with WWTP 5 (ρ = 0.84) likely reflects differences in case variability rather than absolute burden. WWTP 3 exhibited lower day-to-day variance in case incidence during the study period, reducing the discriminatory power of rank-based correlation metrics even when directional trends were broadly consistent.

Hill et al. (2025) compared eight WBE-based *R*_*t*_ estimation approaches and found that methodological choices substantially influenced estimates. The present study does not seek to advance a competing method but rather to assess whether a relatively simple GLMM framework applied to routine twice-weekly sampling can yield transmission trend indicators concordant with case-based surveillance at the sewershed level.

Future research may benefit from evaluating how wastewater-based surveillance scales to larger geographic units, such as counties or clusters of wastewater treatment plants, and whether multiple facilities within a region capture consistent spatiotemporal transmission patterns. Assessing how wastewater-derived indicators can be aggregated without obscuring local dynamics would inform their use in regional surveillance frameworks. As wastewater surveillance continues to be integrated into public health practice, clarifying its role next to the traditional epidemiologic data will be essential for understanding how population-level signals can complement, rather than replace, case-based reporting systems.

## Data Availability

Data is available upon request from Georgia Department of Public Health

https://dph.georgia.gov/phip-data-request

## Acknowledgments

We thank the Georgia Department of Public Health for providing access to sewershed-level COVID-19 case data, and the participating wastewater treatment plant operators and university laboratory partners for sample collection and analysis.

## Data Availability

Code used for this analysis is available at: https://github.com/sethmund/wastewater_glmm. Wastewater and case surveillance data are available upon request from the Georgia Department of Public Health subject to data use agreements.

## Appendix

## References

1. Silk BJ. COVID-19 Surveillance After Expiration of the Public Health Emergency Declaration—United States, May 11, 2023. Morbidity and Mortality Weekly Report. 2023;72:e1.

2. Huisman JS, et al Wastewater-Based Estimation of the Effective Reproductive Number of SARS-CoV-2. Environmental Health Perspectives. 2022;130(5):057011.

3. Zheng X, et al Wastewater Surveillance Provides Spatiotemporal SARS-CoV-2 Infection Dynamics. Engineering. 2024; 40(9) pages 70–77,

4. Safford H, et al Wastewater-Based Epidemiology for COVID-19: Handling qPCR Nondetects and Comparing Spatially Granular Wastewater and Clinical Data Trends. ACS ES&T Water. 2022;2(11):2114–24.

5. Nourbakhsh S, et al A Wastewater-Based Epidemic Model for SARS-CoV-2 with Application to Three Canadian Cities. Epidemics. 2022;39:100560.

6. Hill A, et al Comparison of methods for estimating the effective reproduction number from wastewater data. Epidemics. 2025;52:100839.

7. Ravuri S, et al Estimating effective reproduction numbers using wastewater data from multiple sewersheds for SARS-CoV-2 in California counties. Epidemics. 2025;50:100803.

8. Ungar L. Pandemic Gets Tougher to Track as COVID Testing Plunges. Associated Press. 2022.

9. Lott MEJ, et al Direct Wastewater Extraction as a Simple and Effective Method for SARS-CoV-2 Surveillance and COVID-19 Community-Level Monitoring. FEMS Microbes. 2023;4:xtad004.

10. Sablon O, et al Nanotrap® KingFisher Concentration/Extraction & MagMAX KingFisher Extraction. protocols.io. 2022.

11. Antkiewicz DS, et al Wastewater-Based Protocols for SARS-CoV-2: Insights into Virus Concentration, Extraction, and Quantitation Methods from Two Years of Public Health Surveillance. Environmental Science: Water Research & Technology. 2024;10(8):1766–84.

12. Miyazawa S, et al Wastewater-Based Reproduction Numbers and Projections of COVID-19 Cases in Three Areas in Japan, November 2021 to December 2022. Eurosurveillance. 2024;29(8):2300277.

13. Narci R, et al Inference for Partially Observed Epidemic Dynamics Guided by Kalman Filtering Techniques. Computational Statistics & Data Analysis. 2021;164:107319.

14. Daza-Torres ML, et al A Mixed-Effects Model to Predict COVID-19 Hospitalizations Using Wastewater Surveillance. Journal of Environmental Chemical Engineering. 2024;12(2):112485.

15. de Graaf M, et al Capturing the SARS-CoV-2 Infection Pyramid Within the Municipality of Rotterdam Using Longitudinal Sewage Surveillance. The Science of the Total Environment. 2023;883:163599.

16. Cori A, et al A New Framework and Software to Estimate Time-Varying Reproduction Numbers During Epidemics. American Journal of Epidemiology. 2013;178(9):1505–12.

17. Nash RK, et al Estimating the Epidemic Reproduction Number from Temporally Aggregated Incidence Data: A Statistical Modelling Approach and Software Tool. PLOS Computational Biology. 2023;19(8):e1011439.

18. Kremer C, et al Serial Intervals for SARS-CoV-2 Omicron and Delta Variants, Belgium, November 19-December 31, 2021. Emerging Infectious Diseases. 2022;28(8):1699–1702.

19. Wannigama DL, et al COVID-19 monitoring with sparse sampling of sewered and non-sewered wastewater in urban and rural communities. iScience. 2023;26(7):107019.

20. Rezaeitavabe F, et al Beyond Linear Regression: Modelling COVID-19 Clinical Cases with Wastewater Surveillance of SARS-CoV-2 for the City of Athens and Ohio University Campus. Science of The Total Environment. 2024;912:169028.

21. Deng Y, et al Ratio of Infections to COVID-19 Cases and Hospitalizations in the United States based on SARS-CoV-2 Seroprevalence Data, September 2021-February 2022. Open Forum Infectious Diseases. 2025;12(1):ofae719.

